# Emergent strategies for climate protection among German hospitals — A survey of hospital administrative leaders

**DOI:** 10.1101/2024.10.13.24315412

**Authors:** Lara Schmidt, Sabine Bohnet-Joschko

## Abstract

Hospitals emit large amounts of greenhouse gas emissions during healthcare delivery due to their extensive resource utilization and substantial waste generation. By implementing climate actions, hospitals can significantly contribute to climate protection in healthcare. This paper delves into the climate protection efforts of German hospitals, with a specific focus on the emergent strategies of hospital administrative leaders and the employee engagement within the framework of Stakeholder Theory. The investigation is based on primary data from an online survey of hospital administrative leaders in German hospitals. Employing a hierarchical cluster analysis, the study identifies four distinct clusters of hospitals. These clusters vary significantly in their organization and communication strategies regarding climate protection and allocation of responsibilities, indicating that German hospitals prioritize climate protection to varying degrees. The findings suggest that employee engagement depends on how hospital administrative leaders organize and communicate the topic of climate protection in their institutions. The study underscores the importance of strategic leadership for climate protection in hospitals.

## Introduction

Climate change represents one of the greatest challenges to humankind and significantly impacts both physical and mental health [1]. Examples include heat-related illnesses and deaths due to dehydration, heat stroke, and cardiovascular diseases [2]. Climate-related impacts are projected to increase in intensity throughout this century, threatening to disrupt the ability of healthcare systems to provide quality care [3]. Furthermore, healthcare delivery has also been shown to contribute to climate change [4]. The modern healthcare sector accounts for approximately 4 – 5 % of global greenhouse gas (GHG) emissions [3, 5]. Specifically, hospitals emit large amounts of GHG emissions through their medical care and associated administrative activities [6], utilizing numerous resources and producing various wastes [7]. By taking action to reduce GHG emissions, hospitals can make a significant and ongoing contribution to climate protection [8]. Fundamental climate actions such as strategies for energy efficiency, waste recycling, and low-carbon procurement can be mentioned as steps towards improving the climate, as numerous examples show [9–11].

Implementing climate actions will continue to be necessary as climate protection becomes increasingly important in public health and health policy debates [12]. The increased focus is also driven by global declarations such as the Paris Agreement, as the first universal and legally binding global climate agreement enacted in 2016 [13], or the European Green Deal enacted in 2021 [14]. In addition to political agreements, non-profit organizations are supporting the transformation of the healthcare sector towards greater sustainability. Internationally, the non-profit organization Health Care Without Harm initiated the Global Green and Healthy Hospitals (GGHH) network in 2011. This network, recognized by the World Health Organization, provides a comprehensive framework consisting of ten interconnected objectives for hospitals to promote greater sustainability and environmental health, including:

1. Leadership as a superordinate goal, essential at all levels to promote green and healthy hospitals,
2. Substitution of harmful chemicals with safer alternatives,
3. Reduction, treatment, and safe disposal of medical and non-medical waste,
4. Promotion of energy efficiency and clean, renewable energy generation,
5. Reduction of water consumption and provision of potable water,
6. Improvement of transport strategies for patients and employees,
7. Reduction of food waste and promotion of healthy, locally and sustainably produced food,
8. Reduction of pharmaceutical pollution and prescription of safer pharmaceuticals,
9. Green buildings to develop greener and healthier hospital design and construction,
10. Purchase of sustainable products and materials [15, 16].

With a view to these objectives, current research often addresses energy [17, 18], waste [19, 20], and transportation [21, 22], while areas such as chemicals, water, food, pharmaceuticals, buildings, and purchasing are less investigated in comparison. No study has yet focused on hospitalś administrative leadership and their strategies for climate protection [23]. However, hospitals’ administrative leadership will be in a pole position to plan and implement climate actions and create a fundamental change toward an environmentally sustainable corporate culture through communication and employee engagement [24].

As a global network with more than 1.900 members, GGHH is represented in more than 86 countries [5]. The network includes healthcare organizations from countries such as the United Kingdom, with 38 members, and Australia, with 156 members [25], which are already pursuing national strategies with ambitious targets for climate protection in the healthcare sector [11, 22, 26], while German healthcare organizations are less present with a total of 18 members.

The German healthcare system is often regarded as one of the best in the world, providing universal health insurance coverage and access to a comprehensive range of services with comparatively low cost-sharing requirements. Inpatient care is of great importance, with a dense network of hospitals comprising a mix of public, private not-for-profit, and private for-profit institutions [27]. It includes more than 1.800 hospitals with different bed capacities and levels of care [28], caring for around 17 million cases per year [29]. With European regulations such as the Sustainability Reporting Directive (CSRD) coming into force [30], the vast majority of German hospitals are obliged to include sustainability as an integral part of their corporate strategy in the future [31].

In this context, our study explores the formation of German hospitalś strategic perspectives on climate protection, focusing on hospital administrative leadership and communication. The research question is:

“How are German hospitals strategically positioned to contribute to climate protection?“

### Theoretical perspxective

Companies across various sectors increasingly strive to consider environmental and social aspects within their business activities [32, 33]. In this way, they are responding to the social and political pressure exerted by various stakeholders [34], which has particularly increased with the introduction of CSRD in 2023 [30]. Central to this discourse, the Stakeholder Theory [35] is a framework often used to explain a firm’s sustainability-related behavior [36]. Since its inception [35], Stakeholder Theory has developed into a large and diverse branch of research that comprehensively addresses almost all management sub-disciplines [37].

Stakeholder Theory advocates a management approach that emphasizes the need to consider the interests and needs of all stakeholders, including shareholders, employees, customers, and suppliers [38, 39], who can affect or be affected by the organizatiońs activities [35, 40]. It emphasizes the importance of continuous stakeholder engagement to align stakeholder needs with organizational goals [41, 42] and as a strategic variable to achieve long-term and sustainable business success by strengthening stakeholder cooperation and trust [35, 37, 43].

In the context of hospital administrative leadership’s implementation of climate protection strategies, employees play a crucial and active role in achieving sustainability outcomes. Employees are at the forefront of the organization, contributing innovative ideas, providing solutions to problems, and implementing climate actions [44]. Their involvement in shaping initiatives contributes to broader ecological sustainability performance, enhancing the acceptance of changes within the corporate culture [45]. Therefore, access to information, support, and communication through the administrative leadership is essential for employees to propose initiatives and participate in climate protection [46]. The participatory process of addressing their interests and engagement is essential for strategic alignment in climate protection [47].

In this context, hospital administrative leaders need to find ways to meet their employeeś various interests and needs [35, 43] and simultaneously align these interests through organization and communication [37]. In their pioneering role, administrative leaders can motivate employees to contribute to climate protection [48, 49].

To understand which strategies German hospitals adopt to contribute to climate protection, we examine the administrative leadership regarding their organization and communication and, in this context, the role of employees in implementing climate actions.

## Methodology

A quantitative research methodology was employed to collect numerical data from hospital administrative leaders.

### Sample and data collection

A cross-sectional survey design was utilized for this research. Surveys were conducted across public, private non-profit, and private for-profit hospitals in Germany. Day-care hospitals and psychosomatic clinics were excluded. The studýs sample comprised hospital administrative leaders from 1.169 hospitals. Data was collected between December 12, 2022, and March 19, 2023. Administrative leaders were identified through the hospitaĺs website and quality reports. Survey invitations and a brief introduction of the study were disseminated via email and LinkedIn. In instances where contact information was unavailable, the hospitaĺs email address for general inquiries was used, with a request to forward the survey to the hospital administrative leader. After six weeks, a reminder was sent to encourage participation in the survey.

### Ethics statement

Participation in the online survey was voluntary and did not represent any risk, discomfort or intervention to hospital administrative leaders. The study design and its purpose were explained, contact information were provided to ask any questions related to study. Participants were fully informed about their rights and withdrawal mechanism and gave written consent. Only non-sensitive and professional information was collected. The collected data were kept anonymous and cannot be traced back.

### Questionnaire design

The surveýs design was meticulously crafted, drawing upon an extensive literature review that delved into hospitals’ contributions to climate protection [23, 50]. Before the start of the survey, participants were informed about its duration, data anonymity, and compliance with data protection regulations, which they acknowledged by checking a box. This served as written consent, ensuring that participants were aware of the purpose of the study, how the data would be handled, and their rights before proceeding. The questionnaire was presented in German and comprised six sections:

A. **Socio-demographic data of respondents:** This section captured age, gender, and working experience in hospital administrative leadership.
B. **Characteristics of hospitals**: This section gathered information on the hospitaĺs size, ownership, and level of care.
C. **Climate action implementation:** Focused on collecting data regarding the extent to which and in which areas hospitals have already implemented climate actions and collected performance indicators.
D. **Climate protection initiatives and projects:** Included questions about the hospitaĺs employeeś engagement in adopting climate protection initiatives.
E. **Organization:** Explored the hospitalś organization concerning climate protection, including strategic planning, requirements for suppliers, and the allocation of human resources for coordination and management.
F. **Communication:** Investigated the communication practices of hospital administrative leadership regarding climate protection, including communication about carbon footprint performance indicators, opportunities to contribute ideas for implementing climate actions, and regular meetings for climate protection.

Questions in sections two to five were formatted to allow binary responses (yes/no) and ratings on a Likert scale, ranging from 1 (strongly disagree) to 7 (strongly agree). A Likert scale is predicated on the assumption that personal assessments can be quantitatively measured [51] and was used to analyze the distribution of responses and to group the hospitals.

Pretests were performed with five hospital administrative leaders to evaluate the questionnairés practicability, completeness, and comprehensibility. These tests were also instrumental in identifying and rectifying any potential errors within the online survey. As a result, some questions were restructured and rephrased. The questionnaire was designed using an online survey tool (LimeSurvey version 6.4.1).

### Data analysis

Upon completion of the survey, the data set of 288 individual responses was meticulously screened to ensure the validity of the responses, identify missing values, and recognize outliers [52]. Eighty-three responses were removed as they were not sufficiently completed for our analysis. The data set was transferred to IBM SPSS Statistics version 22, cleaned, and further analyzed. To provide a comprehensive overview, the demographic characteristics of the respondents and the hospitals characteristics were described using descriptive statistics.

### Cluster analysis

We conducted a cluster analysis with inferential tests. The goal was to identify a grouping wherein the characteristics within each cluster are similar yet distinct across clusters [53]. We applied a hierarchical algorithm based on Ward’s method, which is known to be appropriate for small data sets [54], to distinguish homogeneous clusters reflective of hospital climate protection strategies in organization and communication. To identify and, if necessary, exclude outliers, the single-linkage method (nearest neighbor) was employed [55]. The number of clusters was identified using the elbow method, which provides visual support for the cluster decision [56, 57]. Based on this analysis, a four-cluster solution was chosen. Subsequently, we empirically examined the four clusters to compare the differences in means regarding their strategies for climate actions. The Bonferroni post-hoc test confirmed that all variables significantly contribute to the differentiation of the four clusters.

### Inferential statistics

Consistent with other studies [58, 59], we utilized a multiway MANOVA to ascertain significant differences among the contributions to climate protection across the four clusters. MANOVA is a well-established statistical method used to explore the relationships between several categorical independent variables and two or more metric dependent variables [53].

We conducted a chi-square test to assess differences in hospital characteristics. This non-parametric method, chosen for its ability to compare proportions between two or more groups and to test the null hypothesis of independence [60], has practical implications for understanding the distribution of climate actions across the clusters. The MANOVA was also utilized to evaluate the outcomes related to climate actions.

## Results

### Sample characteristics

Our final sample included 205 respondents, with a majority being male (81.5 %). The median age was between 50 and 59 years. Seventy-four respondents (36 %) reported holding an administrative leadership position in multiple hospitals, boosting our study’s implications beyond 205 hospitals within Germany. The respondents’ median work experience was 10 – 14 years (see Table 1).

**Table 1.** Socio-demographic characteristics of the respondents (n=205).

| Characteristics | Sample (%) |
| --- | --- |
| **Age** |  |
| Under 30 | 7.32 % |
| 30 – 39 | 19.02 % |
| 40 – 49 | 23.41 % |
| 50 – 59 | 35.12 % |
| Over 60 | 15.12 % |
| **Gender** |  |
| Male | 81.46 % |
| Female | 18.05 % |
| Diverse | 0.49 % |

| **Administrative leadership position in multiple hospitals** |  |
| --- | --- |
| Yes | 36.10 % |
| No | 63.90 % |
| **Work experience as a hospital administrative leader** |  |
| Less than 5 years | 20.00 % |
| 5 – 9 years | 17.56 % |
| 10 – 14 years | 18.05 % |
| 15 – 20 years | 18.05 % |
| More than 20 years | 26.34 % |

As outlined in Table 2, hospital characteristics reveal a median range of 300 – 499 beds per hospital. Approximately 39.02 % of the surveyed hospitals provide primary and standard care. Hospital ownership was evenly distributed. Hospital administrative leaders from single hospitals (40 %) primarily participated in the survey.

**Table 2.**
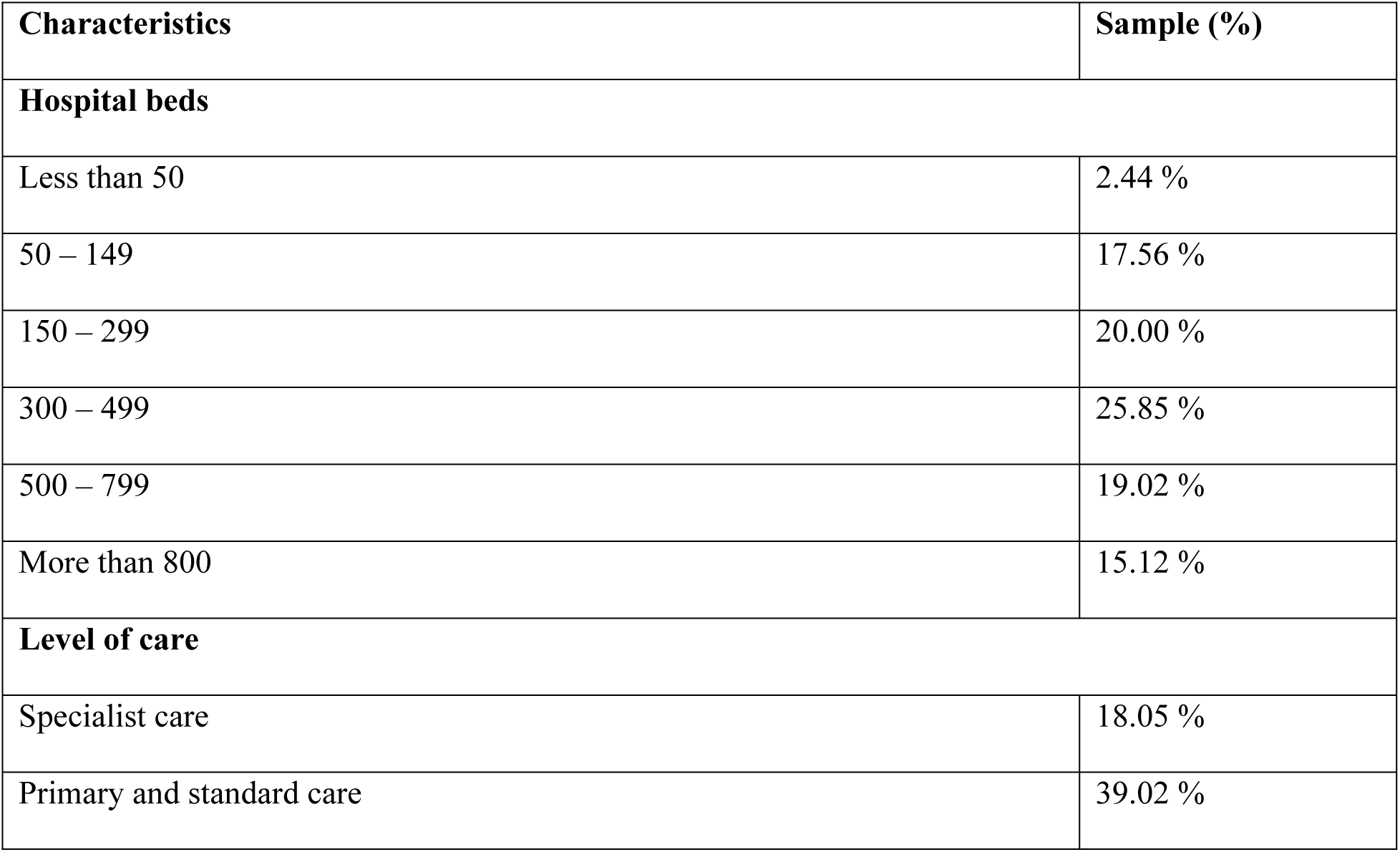
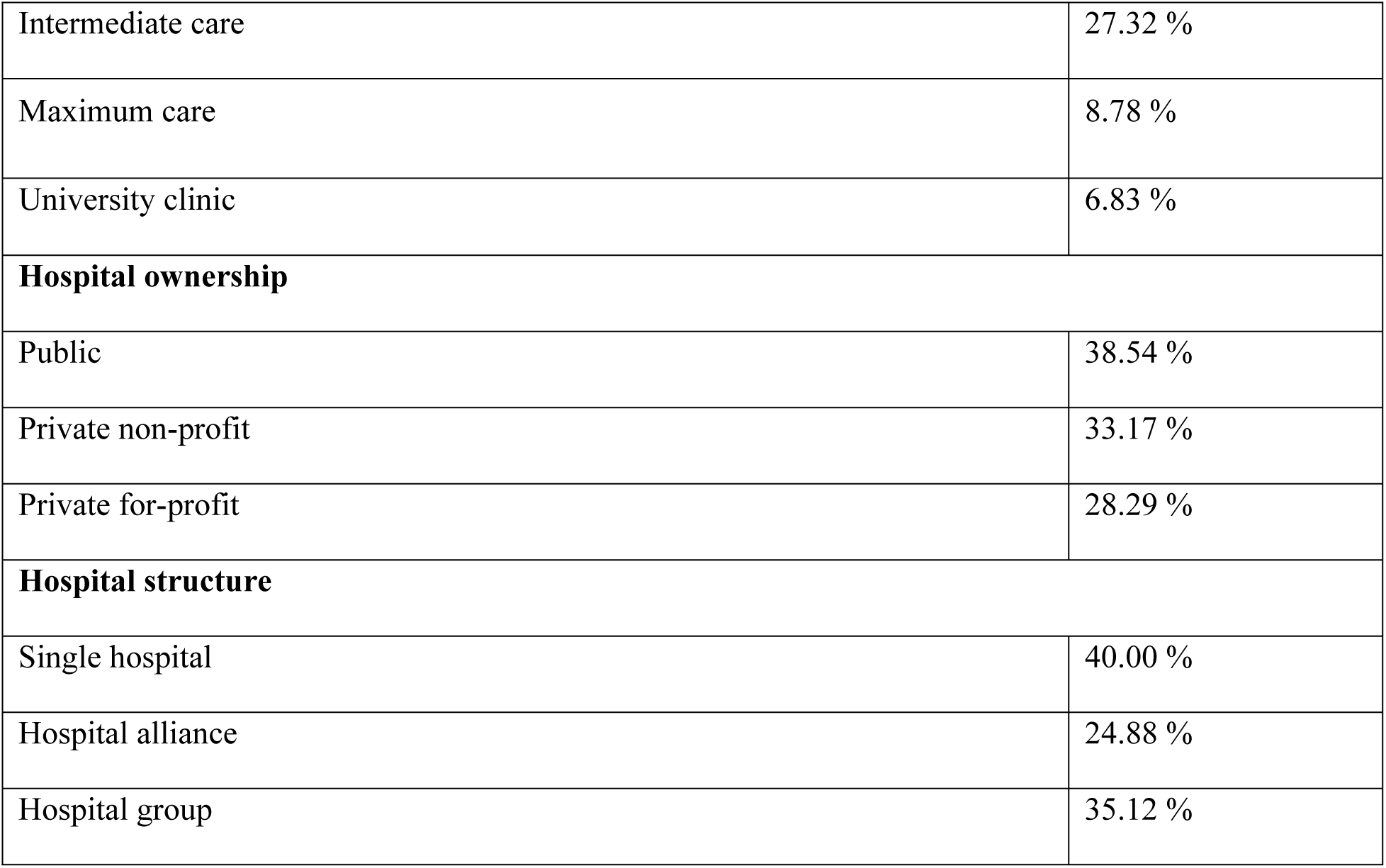
Hospital characteristics (n=205).

| **Characteristics** | **Sample (%)** |
| --- | --- |
| **Hospital beds** |  |
| Less than 50 | 2.44 % |
| 50 – 149 | 17.56 % |
| 150 – 299 | 20.00 % |
| 300 – 499 | 25.85 % |
| 500 – 799 | 19.02 % |
| More than 800 | 15.12 % |
| **Level of care** |  |
| Specialist care | 18.05 % |
| Primary and standard care | 39.02 % |
| Intermediate care | 27.32 % |
| Maximum care | 8.78 % |
| University clinic | 6.83 % |
| **Hospital ownership** |  |
| Public | 38.54 % |
| Private non-profit | 33.17 % |
| Private for-profit | 28.29 % |
| **Hospital structure** |  |
| Single hospital | 40.00 % |
| Hospital alliance | 24.88 % |
| Hospital group | 35.12 % |

The majority of hospitals represented in our study have implemented climate actions in the areas of energy and buildings. Actions were often implemented in hospital engineering, while medical departments were rarely affected. Most hospitals have not set up a sustainability committee or a cost center. The driving force behind the implementation of actions are technical employees, followed by the administrative employees, while the medical employees seem to show little engagement.

### Clustering emergent climate protection strategies in German hospitals

Cluster analysis identified four distinct groups of emergent strategies for climate protection in German hospitals. The clusters, each with its unique characteristics, are described in Table 3. According to the Bonferonni post-hoc test, all selected cluster variables significantly contribute to cluster separation. The results of the MANOVA indicate statistically significant differences among the mean values (mv) for each cluster, as evidenced by p-values for the F-statistic test being less than 0.05. Significant differences include hospital characteristics, the implementation of climate actions, and climate protection initiatives and projects.

**Table 3.**
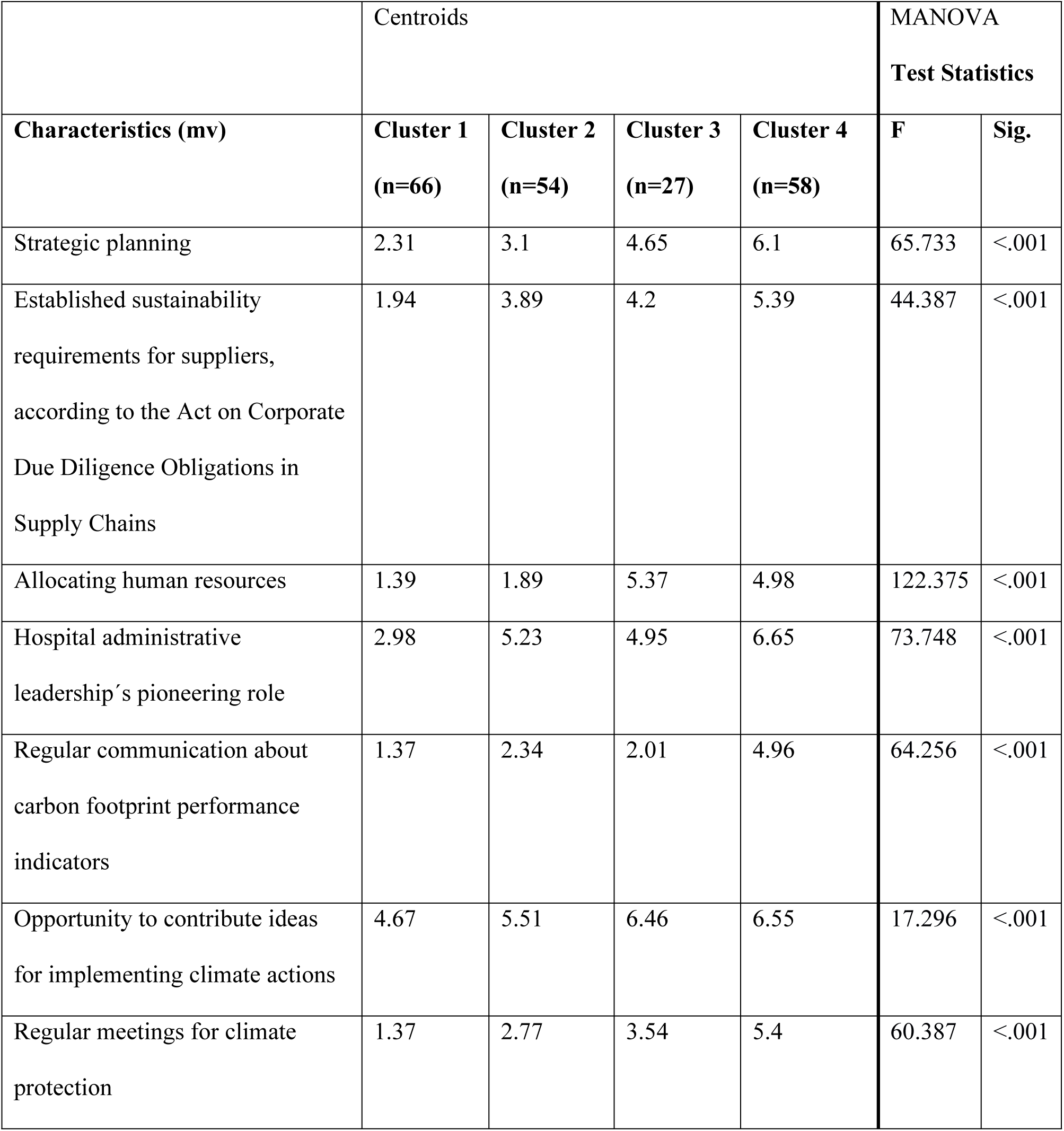
Results of MANOVA.

| Characteristics (mv) | Centroids |  |  |  | MANOVA |  |
| --- | --- | --- | --- | --- | --- | --- |
|  | Cluster 1<br>(n=66) | Cluster 2<br>(n=54) | Cluster 3<br>(n=27) | Cluster 4<br>(n=58) | F | Sig. |
| Strategic planning | 2.31 | 3.1 | 4.65 | 6.1 | 65.733 | <.001 |
| Established sustainability requirements for suppliers, according to the Act on Corporate Due Diligence Obligations in Supply Chains | 1.94 | 3.89 | 4.2 | 5.39 | 44.387 | <.001 |
| Allocating human resources | 1.39 | 1.89 | 5.37 | 4.98 | 122.375 | <.001 |
| Hospital administrative leadership's pioneering role | 2.98 | 5.23 | 4.95 | 6.65 | 73.748 | <.001 |
| Regular communication about carbon footprint performance indicators | 1.37 | 2.34 | 2.01 | 4.96 | 64.256 | <.001 |
| Opportunity to contribute ideas for implementing climate actions | 4.67 | 5.51 | 6.46 | 6.55 | 17.296 | <.001 |
| Regular meetings for climate protection | 1.37 | 2.77 | 3.54 | 5.4 | 60.387 | <.001 |

A radar chart for the clusters enabled in-depth profiling of emergent strategies for climate protection (see Fig. 1). We labeled the first cluster “Aspiring Novices”, as it comprises hospitals with little leadership focus on climate protection, all the same allowing their employees to contribute ideas for climate actions. The second cluster comprises hospitals discovering climate protection and is labeled “Participative Explorers”. A notable feature of this cluster is the dual focus on employeeś ideas and administrative leadership’s pioneering role. We labeled the third cluster “Invested Practitioners”. This cluster not only encourages employee contributions and administrative leadership’s pioneering role but also stands out for allocating additional human resources to facilitate the execution of climate-related actions. The best climate protection strategy characterizes Cluster 4, labeled as “Well-equipped Experts”. It shares the previous clusterś emphasis on employeeś involvement and administrative leadership’s pioneering role but further distinguishes itself through comprehensive strategic planning for climate protection.

**Fig. 1.**
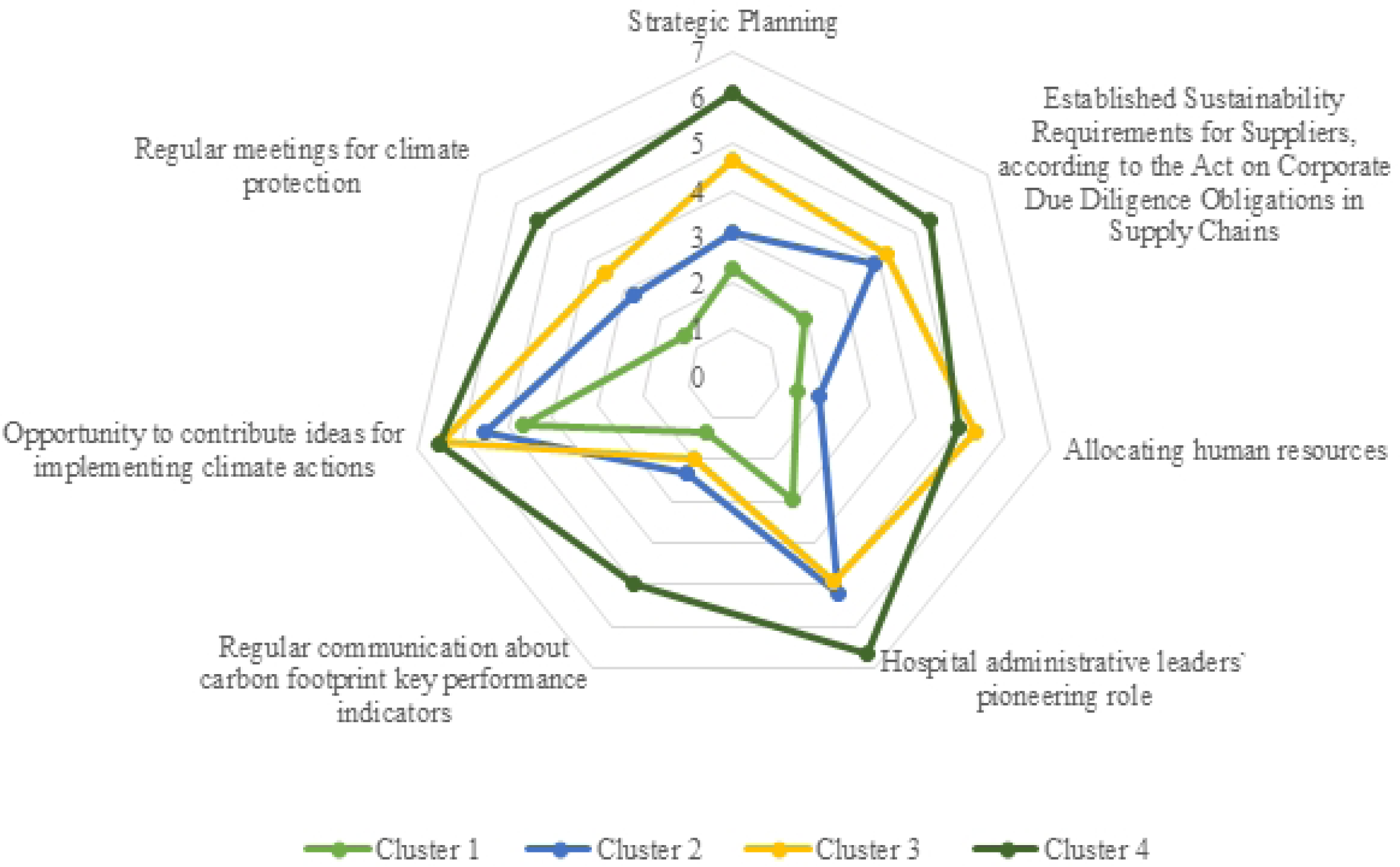
Centroids.

Our research has revealed significant differences in several aspects of climate action implementation. These aspects include the implementation of climate actions, the hospital areas and departments where these actions were implemented, the allocation of responsibilities, the establishment of sustainability committees, and the responsibility among the employees (detailed information in S1 Tables).

## Cluster 1: Aspiring Novices

Comprising 32.20 % of the sample, hospitals in Cluster 1 possess between 50 and 799 beds and are in public ownership. In particular, they are single hospitals that provide primary and standard care, while no university hospital is represented. This group displays a minor engagement with climate protection, exhibiting minimal strategic planning and implementation of climate actions (mean value (mv) = 3.74), primarily implemented in the area of energy (mv = 4.30). Implementing climate actions occurs in hospital engineering (mv = 4.07), while medical departments are marginally affected. Hospital administrative leaders reported unclear responsibilities regarding climate protection (mv = 2.85). 1.52 % of the hospitals in Cluster 1 have established a sustainability committee, and no hospital in this cluster possesses a cost center for implementing climate actions. Administrative leaders also reported collecting key performance indicators for climate protection in energy consumption (mv = 4.36) and water consumption (mv = 4.24). About 56 % of hospitals have taken initiatives and projects for climate protection. While technical employees initiated initiatives and projects (mv = 5.22), hospital administrative leaders estimated limited climate protection efforts by medical employees. In the future, the technical employees (mv = 5.94) and the hospital administrative leadership (mv = 5.87) should expand their responsibility for climate protection while no additional engagement is expected from medical employees.

### Cluster 2: Participative Explorers

Hospitals in Cluster 2 (26.34 %) possess a medium number of beds and are mainly privately owned. In particular, they are part of a hospital group and provide primary and standard care. This group shows a moderate commitment to climate protection with moderate strategic planning efforts. Although the implementation of climate actions (mv = 4.63) is higher than in Cluster 1, it is in the lower range compared to the other clusters. The implementation of climate actions (mv = 4.63) focuses on energy (mv = 4.99) and food (mv = 4.79). Hospital administrative leaders reported implementing climate actions in hospital engineering (mv = 5.13) and food supply (mv = 4.61), while, similar to Cluster 1, the medical departments are slightly affected. Responsibilities for climate protection are significantly higher than in Cluster 1 but remain at a medium level in group comparison (mv = 4.11). In this cluster, 25.93 % of hospitals have established a sustainability committee, and 3.70 % possess a cost center for implementing climate actions. Key performance indicators for climate protection are collected in more areas than in Cluster 1, but least in the consumption quantity of transportation (mv = 3.58) as well as in consumption through procurement (mv = 3.74) and chemical disposal and anesthesia suction (mv = 3.78). About 87 % of the hospital administrative leaders in Cluster 2 reported taking initiatives and projects for climate protection. Technical (mv = 5.12) and, in some cases, administrative employees (mv = 4.66) lead initiatives for climate protection, while medical employees show little initiative. In the future, all professional groups within the hospitals should take more responsibility for climate protection. The primary responsibility will remain with the hospital administrative leadership (mv = 6.11).

### Cluster 3: Invested Practitioners

Hospitals in Cluster 3 (13.17%) possess a medium number of beds and are primarily non-profit in their ownership. In particular, they are part of a hospital alliance and have the highest proportion of specialized care clinics in cluster comparison. They demonstrate a higher commitment to climate protection with a moderate strategic focus. Hospitals in Cluster 3 implement climate actions in almost all areas (mv = 5.68), with a particular focus on energy (mv = 5.87) and the area of pharmaceuticals only being moderately developed (mv = 4.06). In a cluster comparison, hospitals in Cluster 3 implement an above-average number of climate actions in chemicals (mv = 4.41). Unlike previous clusters, hospital administrative leaders reported implementing more climate actions in medical departments, except for the intensive care unit (mv = 3.95). Clearly defined responsibilities for implementing climate actions characterize Cluster 3 (mv=5.66). In this cluster, 33.33 % of hospitals have established a sustainability committee, and 14.81 % possess a cost center for implementing climate actions. Key performance indicators for climate protection are collected in all areas except for the consumption of transportation (mv = 3.84) and consumption through procurement (mv = 3.79). In a cluster comparison, the most critical performance indicators for food disposal are collected in Cluster 3 (mv = 4.88). About 96.30 % of the hospital administrative leaders reported taking initiatives and projects for climate protection. Although technical employees (mv = 5.29) and administrative employees (mv = 4.83), in particular, are driving forward climate protection initiatives, medical employees also make a moderate contribution to climate protection (mv = 4.2). The contribution of the physicians was the highest in cluster comparison (mv = 3.92). In the future, all professional groups should assume more responsibility, with the hospital administrative leaders assigning the most responsibility to employees in functional service in cluster comparison (mv = 4.93).

### Cluster 4: Well-equipped Experts

Cluster 4 (28.29 %) comprises large hospitals with the highest proportion of university hospitals. These hospitals are single hospitals, part of a hospital group, and are in public ownership. They are most actively engaged in climate protection and possess a comprehensive strategic planning. They report extensive climate actions (mv = 6.37) across all areas except chemicals (mv = 3.3), focusing on energy (mv = 5.87), food (mv = 5.4), and buildings (mv = 5.35). Climate actions are actively implemented in all hospital areas and departments. Hospital administrative leaders reported clear responsibilities regarding the implementation of climate actions (mv = 6.43). About 56.90 % of hospitals established a sustainability committee, and 39.66 % reported a cost center for implementing climate actions. They also reported collecting key performance indicators for climate protection in all areas except food supply (mv = 3.83). All hospitals have taken initiatives and projects for climate protection. In cluster comparison, employees’ contribution to initiatives and projects in Cluster 4 is at the highest level, except for medical employees (mv = 3.8). In the future, all professional groups should take more responsibility for the climate. The primary responsibility should remain with the administrative leadership (mv = 6.2) and the technical employees (mv = 6.11).

The hospital structure data for each cluster is provided in Table 4.

**Table 4.** Hospital structure of the four clusters.

| Category | Cluster 1<br>(n=66) | Cluster 2<br>(n=54) | Cluster 3<br>(n=27) | Cluster 4<br>(n=58) |
| --- | --- | --- | --- | --- |
| **Hospital beds** |  |  |  |  |
| Less than 50 | 1.52 % | 5.56 % | 3.70 % | 0.00 % |
| 50 – 49 | 25.76 % | 16.67 % | 14.81 % | 10.34 % |
| 150 – 299 | 18.18 % | 20.37 % | 14.81 % | 24.14 % |
| 300 – 499 | 21.21 % | 29.63 % | 33.33 % | 24.14 % |
| 500 – 799 | 24.24 % | 14.81 % | 18.52 % | 17.24 % |
| More than 800 | 9.09 % | 12.96 % | 14.81 % | 24.14 % |
| **Level of care** |  |  |  |  |
| Specialist care | 22.73 % | 18.52 % | 25.93 % | 8.62 % |
| Primary and standard care | 39.39 % | 44.44 % | 29.63 % | 37.93 % |
| Intermediate care | 25.76 % | 27.78 % | 25.93 % | 29.31 % |
| Maximum care | 12.12 % | 7.41 % | 7.41 % | 6.90 % |
| University clinic | 0.00 % | 1.85 % | 11.11 % | 17.24 % |
| **Hospital ownership** |  |  |  |  |
| Public | 45.45 % | 29.63 % | 29.63 % | 43.10 % |
| Private non-profit | 30.30 % | 24.07 % | 51.85 % | 36.21 % |
| Private for-profit | 24.24 % | 46.30 % | 18.52 % | 20.69 % |
| **Hospital structure** |  |  |  |  |
| Single hospital | 56.06 % | 27.78 % | 25.93 % | 39.66 % |
| Hospital alliance | 22.73 % | 22.22 % | 48.15 % | 18.97 % |
| Hospital group | 21.21 % | 50.00 % | 25.93 % | 41.38 % |

## Discussion

Environmental sustainability is an integral aspect of corporate strategies, and garners increased attention among companies. This trend, well documented in recent literature [61–63], aligns with objectives of enhancing operational efficiency, fostering competitive advantage, and contributing to climate protection efforts [64]. The healthcare sector, in particular, is advancing towards embracing ecological sustainability principles. Academic research on environmental sustainability in hospitals has primarily concentrated on energy efficiency and waste management practices [17]. Little is known about hospitalś administrative leadership strategies for climate protection [23]. This study aimed to scrutinize the strategic position of German hospitals towards climate protection and assess hospitalś contribution to climate protection from the administrative leadership’s perspective. Our research focuses on the organization and communication characteristics of German hospitals and explores different stages in strategy development for climate protection while underscoring the engagement of hospital employees in climate protection initiatives.

We grouped hospitals into four distinct clusters and identified stages of strategic development toward climate protection. Hospitals were categorized into Cluster 1 through 4, while Cluster 1 indicates minimal engagement, whereas Cluster 4 signifies the most proactive attitude. Each cluster with unique characteristics highlights the relevance of administrative leadership’s organization and communication in hospitalś climate protection efforts. The results indicate that a minority of hospitals (28.29 %) attained the highest level of commitment (Cluster 4), while a smaller fraction (13.17 %) was classified into Cluster 3. However, a significant majority (58.54 %) fell into the medium and low engagement categories (Cluster 2 and 1), underscoring a need for more proactive administrative leadership in addressing climate protection. These findings confirm previous research indicating that German hospitals are yet to fully embrace active roles in climate protection [65, 66].

### Characteristics of hospitals

While our analysis showed no differences based on hospital ownership, other structural characteristics proved statistically significant. Specifically, single hospitals demonstrated less commitment to climate protection efforts. In contrast, hospitals affiliated in groups exhibited a markedly higher commitment to climate protection initiatives, including notable efforts to engage employees in these processes. A comparative analysis across four homogenous clusters further highlights this trend, revealing that larger hospitals within groups are better positioned to implement sustainability actions effectively than smaller and single hospitals. This advantage might be attributed to larger institutionś superior resources and organizational framework facilitating more comprehensive and impactful climate protection initiatives. Regulations, such as the CSRD, might be a driver in disclosing the social and environmental risks hospitals face [30]. The anticipated transparent description of GHG emissions will reveal the extent of hospitalś climate impact and the average contributions to climate protection [31].

### Climate action implementation

Although most administrative leaders reported implementing climate actions, there are marked differences in scope. Operations and facility management emerge as common focal points across Clusters 2, 3, and 4 highlighting energy as a central area of operations among respondents. This result is also reflected in the survey of key performance indicators for climate protection. While all hospitals collect data on energy consumption, data collection for chemical disposal, anesthesia suction, and food disposal seems to be of secondary importance. The marginal report of actions in hospital-specific domains such as chemicals or pharmaceuticals illustrates hospital administrative leaderś underestimation of specific GHG sources as reported by previous studies [67]. A significant part of hospital emissions are generated by hospital-specific emission sources. For example, medical supplies and medicines alone are responsible for up to 50 % of overall emissions [68]. Although the bulk of GHG emissions generated during patientś healthcare delivery is indispensable [69], the potential for climate actions in medical departments of German hospitals can be further exploited.

### Climate protection initiatives and projects

We observed distinct variations in allocating responsibilities and human resources for climate action efforts across clusters. Clusters 1 and 2 do not allocate additional human resources for implementing climate actions, whereas Clusters 3 and 4 do. In hospitals without additional human resources, initiatives predominantly originate from technical employees (Cluster 1) and, to a lesser extent, administrative employees (Cluster 2). Hospital medical employees contribute to climate protection, where additional human resources are created, and clear responsibilities exist (Cluster 3 and 4). Assigning overarching responsibility, for example, by creating a sustainability committee, can support the involvement of various employees within the hospital. Previous studies emphasize that employee engagement is crucial in developing and promoting environmental responsibility [70].

### Organization

The clusters show considerable differences in the hospitalś organization regarding climate protection efforts. The implementation of climate actions across various hospital areas and departments critically hinges on the strategic planning undertaken by hospitalś administrative leadership, which is engaged for climate targets in a pioneering role (Cluster 1, 2, 3). Due to challenges such as the shortage of skilled employees and financial performance [71], climate protection is not a strategic priority in many hospitals, except those in Cluster 4. While the Act on Corporate Due Diligence Obligations in Supply Chains was introduced in Germany in 2023 [72], hospitals in Cluster 1 and 2 do not have established sustainability requirements for suppliers. The Act obliges organizations to ensure complete transparency along their supply chain and is intended to ascertain compliance with human rights and environmental obligations. Hospitals are obliged to fulfill their due diligence obligations and to transparently document these in an annual report [72]. Additionally, all respondents reported the pioneering role of hospital administrative leadership as pivotal (except in Cluster 1). Our approach exceeds previous research [70] by identifying emergent strategy stages.

### Communication

Our findings indicate the relevance of employeeś engagement through communication. Communication about climate protection is often associated with greater employee initiatives (Cluster 4), which is why this can be understood as a critical factor for the long-term success and legitimacy of further actions. Hospital stakeholderś awareness of GHG emissions and their sources is imperative for developing and implementing effective strategies for climate protection [67]. Thus, involving and addressing hospital employeeś expectations regarding climate issues is paramount [65, 66]. In contrast to earlier research, suggesting a lack of employeeś influence on climate actions [67], our study indicates that employees across all hospitals can contribute ideas for implementing climate protection initiatives.

In summary, the significant disparities among the clusters underscore that climate protection is still a new topic in the German hospital sector, with many institutions yet to prioritize it. Only hospitals in Cluster 4 exemplify adherence to Stakeholder Theory principles [73, 74], demonstrating a commitment to strategic planning, employee involvement in decision-making processes, assignment of clear responsibilities, and transparent communication regarding climate protection efforts and emissions data. These findings emphasize the interconnectedness of hospital administrative leadership’s influence and employeeś engagement, advocating for continued administrative leadership support of climate protection initiatives [75]. It is crucial that hospital administrative leadership maintains its commitment to climate protection, as changing administrative leadership strategies can cultivate employee awareness, dedication, and actions toward climate protection [76].

To advance strategic objectives toward sustainability in German hospitals, it is crucial to investigate further the reasons behind the varying levels of climate actions across hospitals and to emphasize climate protection as a central leadership issue. Our study emphasizes the central role of hospital administrative leadeŕs strategy and communication and the engagement of employees in climate protection efforts.

## Limitations

This study is the first to analyze the contribution of German hospitals to climate protection from the perspective of their administrative leadership. While extensive in its multifaceted analysis, this study acknowledges certain limitations that open avenues for future research. Due to the current challenges facing German hospitals, such as skill shortages, climate protection is not a top issue for hospital administrative leaders, complicating recruitment efforts for this group. General challenges in recruiting administrative leaders for an online survey can also be observed in other studies among leaders [77, 78]. The usual limitations of an online survey apply [79, 80]. Although the questionnaire was sent exclusively to hospital administrative leaders, its anonymous nature means we cannot guarantee that it was completed solely by our target group. Additionally, comprehension problems on the part of the respondents cannot be recognized and rectified when using an online survey. However, this research approach was best suited to the aim of the study and the chosen target group. Although we have excluded day hospitals and psychiatric facilities, our results show emergent strategies that are applicable to all hospitals and relevant to all healthcare organizations providing medical care. We could not incorporate other stakeholder’ perspectives as we focused on assessing the hospital’s administrative leadership. Exploring the perceptions of other hospital stakeholders could be a promising avenue for further research. The results may only partially reflect the diverse perspectives of the hospital administrative leaders, as interest and awareness of environmental issues beyond legal requirements vary. Those predisposed to climate protection might have been more likely to participate in this study than those less engaged with environmental issues. Additionally, we cannot dismiss the possibility of social desirability bias.

## Conclusion

The urgency of climate change continues to require strategies from the healthcare sector. However, more research on corporate strategies and hospital administrative leadership practices is still needed. To our knowledge, this study represents the first comprehensive analysis of German hospitals’ contribution to climate protection as reported by hospital administrative leaders. We identified four distinct clusters of hospitals associated with various structural characteristics and administrative leaderś strategies regarding climate protection. The results show that large hospitals contribute extensively to climate protection and possess a strategic planning. This can be attributed to the availability of personnel and financial resources, societal pressure to reduce GHG emissions, and the CSRD. Climate actions were mainly implemented in the area of energy. Key performance indicators on energy consumption are collected in all hospitals. Medical departments have, as yet, no significant role in climate protection. Technical and administrative employees are the driving force behind the implementation of climate actions. Clear climate protection strategies are not yet in place in most hospitals. As communication is often associated with greater initiative on the part of employees, it can be accentuated as a crucial indicator for the long-term success and legitimacy of further climate actions in German hospitals. The study highlights the significance of hospital administrative leadership and employee engagement for corporate sustainability and emphasizes the role of sustainable administrative leadership as essential for hospitalś contribution to planetary health.

## Data Availability

All relevant data are within the manuscript and its Supporting Information files.

## Notes

### Competing Interest Statement

The authors have declared no competing interest.

### Funding Statement

The author(s) received no specific funding for this work.

